# Machine Learning Prediction of Surgical Intervention for Small Bowel Obstruction

**DOI:** 10.1101/2021.04.13.21255428

**Authors:** Miles Turpin, Joshua Watson, Matthew Engelhard, Ricardo Henao, David Thompson, Lawrence Carin, Allan Kirk

## Abstract

Small bowel obstruction (SBO) results in >350,000 operations and >$2 billion annual health care expenditures in the US. Prompt, effective identification of patients at high/low surgery risk could improve survival, lower complication rates, and shorten hospitalization lengths. SBO surgery prediction models were developed based on SBO-related encounters in the Duke University Health System between 2013 and 2017. A total of 3,910 encounters among 3,374 unique patients were identified. Performance was assessed in each hour after admission when predicting whether patients will (a) receive surgery within 24 hours, and (b) receive surgery during the current encounter. Potential benefits of model-based discharge were assessed using the incorrect discharge rate and average reduction in hospital stay. Model-based discharge of low-risk patients was projected to reduce the average length of stay among patients not receiving surgery by >60 hours while maintaining an incorrect discharge rate lower than the observed readmission rate (9.3%). AUROC for the 24-hour prediction task increased from 0.644 to 0.779 at 12 and 72 hours post-admission, respectively. Future work will prospectively explore the benefits of model deployment in an inpatient setting.

## INTRODUCTION

Small bowel obstruction (SBO) continues to be one of the most common reasons for inpatient admissions in the US. More than 350,000 SBO operations are performed every year, resulting in 960,000 inpatient days and over $2 billion in health care expenditures.^1^ A recent retrospective national study demonstrated that lysis of adhesions and small bowel resections were two of the seven most common emergency general surgery procedures, and these seven account for the highest admissions, deaths, complications, and inpatient costs of all emergency general surgical procedures.^2^

Three quarters of SBOs are caused by adhesive disease,^3–4^ and in these patients, non-operative management results in resolution 74% of the time.^5^ A number of decision-support tools have been developed to identify patients who can be managed non-operatively versus those requiring surgical intervention, including scoring systems based on radiologic findings, clinical findings, and combinations of the two. Early, accurate risk stratification is needed to mitigate negative outcomes associated with delayed clinical decision-making, including increased morbidity, increased cost, and increased post-operative length of stay.^6–8^ The early identification of the need for surgical intervention and subsequent surgical treatment has demonstrated significant survival benefit, lower incidence of local systemic complications, and shorter hospitalizations.^7^

Although many of these findings have been known for decades, current practice continues to rely on ad hoc management strategies that differ between specialists. Some studies have suggested that management of adhesive-SBO by a primary medical (rather than surgical) team is associated with a higher healthcare utilization and worse perioperative outcomes.^1,9,10^ An objective decision-making tool that updates patients’ risk of requiring surgery as clinical and radiological findings are collected will help to streamline clinical practice and improve outcomes.

The aim of this study was to develop a predictive model capable of assisting clinicians as they assess the need for surgery in patients with SBO. Model development was based on admissions taking place within the Duke University Health System (DUHS) between 2013 and 2015, including readily available laboratory data, nursing collected information, and imaging when available. We hypothesized that this model would effectively stratify patients by their risk of requiring surgery, and that its performance would improve as data accumulated during the encounter. Importantly, we aimed to explore the cost/benefit of reducing the average length of stay for SBO by more promptly discharging patients whose model-predicted risk was low.

## METHODS

### Cohort Identification

Results were based on retrospective data analysis conducted at the Duke University School of Medicine and determined to be exempt from review by the Duke Health Institutional Review Board. All methods were performed in accordance with all relevant guidelines and regulations, including Duke University Health System, Duke Health Institutional Review Board, and Duke University School of Medicine policies. Analyses were executed within the Duke Protected Analytics Computing Environment (PACE), a highly protected virtual network space designed for protected health information. Participant consent was waived by the Duke Health Institutional Review Board due to the minimal risk posed by study procedures and infeasibility of obtaining consent in a large retrospective cohort.

DUHS encounters in an observation window ranging January 1, 2013 – December 31, 2017 were initially identified. Inclusion criteria were (a) age ≥18, and (b) SBO diagnosis present on admission, as determined via retrospective billing data. SBO diagnoses were defined as codes 560.1 (paralytic ileus), 560.8X (other specified intestinal obstruction), or 560.9 (unspecified intestinal obstruction) from the International Classification of Diseases, 9^th^ Revision, Clinical Modification (ICD9-CM). Diagnosis codes 560.2 (intussusception), 560.2 (volvulus), and 560.3X (impaction of intestine) were excluded. Encounters were labeled as ‘Surgery’ or ‘Discharge’ based on the presence or absence of surgery for SBO during the encounter, as defined by the following ICD9-CM procedure codes suggestive of laparoscopic or open intervention: 49320, 49000, 49321, 49002. Other exclusion criteria were (a) over 3 SBO-related encounters in the observation window, and (b) surgery for SBO over 3 weeks after admission.

### Data Preprocessing

For each encounter, a time series of 24 clinical variables was extracted, including vital signs (*e*.*g*., pulse, blood pressure), 11 laboratory results (*e*.*g*., complete blood count, comprehensive metabolic panel), and 8 inputs and outputs (*e*.*g*., intravenous fluid, stool and urine output). Variables were aggregated by hour by taking the mean or sum of all measurements, as appropriate. All variables and missingness rates are presented in Supplementary Tables 1 and 2, respectively.

Missing values were imputed by carrying the last value forward or filling with the median across all encounters when no previous value was available. For each variable, the time since last measurement was included as an additional predictor. Three summary statistics were also calculated per raw variable to capture long-term and short-term trends. Exponential moving averages were calculated for vital signs (6-, 24-, and 72-hour half-life) and laboratory results (12-, 48-, and 144-hour half-life). Exponential moving sums were calculated for inputs and outputs (1-, 6-, and 24-hour half-life).

To incorporate radiological findings, frequently occurring text sequences (1, 2, and 3 words in length) were extracted from radiology notes (*e*.*g*., CT and small bowel follow through studies) associated with both surgical and non-surgical encounters. Text sequences identified as clinically meaningful and interpretable (Supplementary Table 3), as well as indicators for CT and small bowel follow through studies, were used to create predictors indicating (a) appearance in the past 24 hours, and (b) appearance in the encounter up to the current time.

Age, hospital name, and the number of past SBO diagnoses were included as additional static predictors. Hours since admission and time of day were included as additional time-varying predictors. In total, 161 predictors were available for modeling.

### Model Development

A Cox proportional hazards (Cox PH) framework was utilized to predict patients’ relative risk of requiring surgery given the predictor variables available at each time point.^11^ For any time point of interest, this approach allows the model to predict which patients are likely to receive surgery earlier versus later given that they have not yet received surgery by that time. If no SBO surgery took place during the encounter, the time to surgery was right censored for model training. The model was trained to maximize the Cox PH partial likelihood, which quantifies the probability of the observed order of surgery events, including censored events (*e*.*g*., discharges), given the parameters of the model. An elastic net penalty was placed on model parameters to encourage sparsity and reduce overfitting.

The model was implemented and trained in Glmnet for Python with Python 3.5.2.^12^ Training and evaluation took place using 5-fold nested cross-validation in which model performance is evaluated on each of 5 subsets of the data (*i*.*e*., outer folds) after training on the other 4. In nested cross-validation, hyperparameters are selected through a similar procedure, wherein the training data are further partitioned into subsets.^13^ Hyperparameters are then selected to maximize average performance across these subsets (*i*.*e*., inner folds), preventing bias during model selection.^13^ In the current model, hyperparameters included the two elastic net penalties corresponding to L1- and L2-regularization. Stratification was used to ensure the number of encounters resulting in SBO surgery was similar between partitions. All encounters for a given patient were assigned to the same partition(s).

### Statistical Analysis and Performance Measures

Demographic factors and other descriptive statistics were compared between groups by two-tailed Mann-Whitney *U* test for numeric variables, and by chi-square test for categorical variables (see Table 1).

**Table 1:** Demographics and other Descriptive Statistics.

| Variable | Value | Surgery | No surgery | Test Statistic | p-value |
| --- | --- | --- | --- | --- | --- |
| **Encounters** | N (%) | 598 (15.3%) | 3310 (84.7%) | - | - |
| **Age** | Mean ± SD<br>(range) | 61.6 ± 16.6<br>(18-97) | 61.4 ± 16.4<br>(18-109) | $U=9.9e5$ | 0.68 |
| **Length of Stay** | Mean ± SD<br>(range) | 13.3 ± 11.3<br>(0.76-89.2) | 7.6 ± 9.5<br>(0.11-252.8) | $U=1.4e6$ | 0 |
| **Weight** | Mean ± SD<br>(range) | 171.5 ± 46.9<br>(70-433.4) | 174.0 ± 51.7<br>(48.3-456.3) | $U=9.3e5$ | 0.03 |
| **Sex** | *Female*, N (%) | 340 (56.9%) | 1746 (52.7%) | $\chi^2=3.26$ | 0.07 |
|  | *Male*, N (%) | 258 (43.1%) | 1564 (47.3%) |  |  |
| **Location** | *Duke University Hospital*, N (%) | 105 (17.6%) | 531 (16.0%) | $\chi^2=3.75$ | 0.15 |
|  | *Duke Raleigh Hospital*, N (%) | 160 (26.8%) | 796 (24.0%) |  |  |
|  | *Duke Regional Hospital*, N (%) | 333 (55.7%) | 1983 (59.9%) |  |  |
| **Ethnicity** | *Hispanic or Latino*, N (%) | 14 (2.3%) | 69 (2.1%) | $\chi^2=0.16$ | 0.99 |
|  | *Not Hispanic/Latino*, N (%) | 573 (95.8%) | 3180 (96.1%) |  |  |
|  | *Not Reported/Declined*, N (%) | 11 (1.8%) | 61 (1.8%) |  |  |
| **Race** | *Two or more races*, N (%) | 7 (1.2%) | 29 (0.9%) | $\chi^2=12.82$ | 0.08 |
|  | *American Indian or Alaskan Native*, N (%) | 7 (1.2%) | 22 (0.7%) |  |  |
|  | *Asian*, N (%) | 15 (2.5%) | 38 (1.1%) |  |  |
|  | *Black or African American*, N (%) | 179 (29.9%) | 1098 (33.2%) |  |  |
|  | *Caucasian/White*, N (%) | 375 (62.7%) | 2022 (61.1%) |  |  |
|  | *Native Hawaiian or Other Pacific Islander*, N (%) | 0 (0.0%) | 2 (0.1%) |  |  |
|  | *Not Reported/Declined*, N (%) | 3 (0.5%) | 34 (1.0%) |  |  |
|  | *Other*, N (%) | 12 (2.0%) | 65 (2.0%) |  |  |
| **Insurance** | *Commercial*, N (%) | 9 (1.6%) | 38 (1.2%) | $\chi^2=15.8$ | 0.11 |
|  | *Managed Care*, N (%) | 60 (10.4%) | 400 (12.4%) |  |  |
|  | *Medicaid Pending*, N (%) | 0 (0.0%) | 2 (0.1%) |  |  |
|  | *Medicare*, N (%) | 253 (43.7%) | 1431 (44.3%) |  |  |
|  | *Medicare Advantage*, N (%) | 89 (15.4%) | 409 (12.7%) |  |  |
|  | *NC Blue Cross*, N (%) | 84 (14.5%) | 397 (12.3%) |  |  |
|  | *NC Medicaid*, N (%) | 36 (6.2%) | 242 (7.5%) |  |  |
|  | *OOS Blue Cross*, N (%) | 34 (5.9%) | 213 (6.6%) |  |  |
|  | *OOS Medicaid*, N (%) | 5 (0.9%) | 9 (0.3%) |  |  |
|  | *Other Government*, N (%) | 3 (0.5%) | 25 (0.8%) |  |  |
|  | *Special Programs*, N (%) | 6 (1.0%) | 61 (1.9%) |  |  |

The model was evaluated based on its ability to predict (a) whether patients will require surgery within 24 hours, and (b) whether patients will require surgery at any time during the encounter. A 24-hour prediction window was chosen because it is long enough to allow surgeons to intervene, but short enough to allow the model to effectively identify patients whose condition is deteriorating.

Performance for these binary prediction tasks was quantified via the receiver operating characteristic (ROC) curve, which depicts the tradeoff between true positive rate (TPR; *i*.*e*., sensitivity) and false positive rate (FPR; *i*.*e*., 1 – specificity), as well as the area under the ROC curve (AUROC), which measures performance across all TPR-FPR pairs.^14^ The AUROC may also be interpreted as the model’s effectiveness in ranking encounters resulting in surgery as higher risk than encounters not resulting in surgery. A final evaluation measure, the concordance index (C-index), measures the model’s effectiveness in (a) ranking encounters resulting in surgery as higher risk than those not resulting in surgery, and (b) correctly ranking encounters resulting in surgery by the time elapsed between admission and surgery.^15^

Aggregate performance across all time points would not be meaningful due to differences in length of stay between individual encounters and the average length of stay in ‘Surgery’ versus ‘Discharge’ groups. Further, very few laboratory results and inputs and outputs are available within the first few hours of admission. Instead, performance was calculated at specific time points after admission to provide information directly relevant to a prospective deployment. Performance has also been evaluated by the time remaining until surgery or discharge to examine how performance changes when approaching these endpoints.

An additional “early discharge task” was explored to evaluate the model’s effectiveness in discharging low-risk patients. In this task, patients are discharged whenever their model-predicted risk falls below a fixed threshold. Early discharge performance may then be quantified across a range of thresholds using task-specific metrics. These include (a) the *discharge predictive value* (*i*.*e*., positive predictive value for the discharge task), which quantifies the proportion of patients identified by the model for early discharge who did not later require surgery; (b) the *true discharge rate* (*i*.*e*., sensitivity for the discharge task), which quantifies the proportion of patients eventually discharged who were correctly identified; (c) the *incorrect discharge rate* (*i*.*e*., false positive rate for the discharge task), which quantifies the proportion of patients requiring surgery who were incorrectly identified for discharge; and (d) the *average reduction in hospital stay* among patients who did not receive surgery.

To compare model-based discharge with clinician performance, the incorrect discharge rate observed in our cohort was estimated by defining a false negative as a case where a patient requiring surgery had been readmitted for SBO fewer than three days after a previous discharge without surgery.

## RESULTS

### Description of Participants

A total of 7191 SBO-related encounters were identified between January 1, 2013 and December 31, 2017. After applying exclusion criteria, 3,910 encounters among 3,374 unique patients remained for analysis. Of these patients, 13.7% were readmitted at least once during the observation window (*i*.*e*., 2013-2017) with an SBO diagnosis on admission. SBO-related surgery was identified in 606 of the encounters (‘Surgery’; 15.5%), whereas no surgery was identified in 3304 (‘Discharge’; 84.5%). Stepwise application of exclusion criteria is shown in Figure 1. Demographics, length of stay, and other descriptive statistics are presented in Table 1. SBO surgery most commonly occurred in the first 24 hours after admission (median=25 hours). Patients not receiving surgery were most commonly discharged between 4 and 5 days after admission (median=4.7 days). Detailed encounter endpoints are presented in Figure 2.

**Figure 1:**
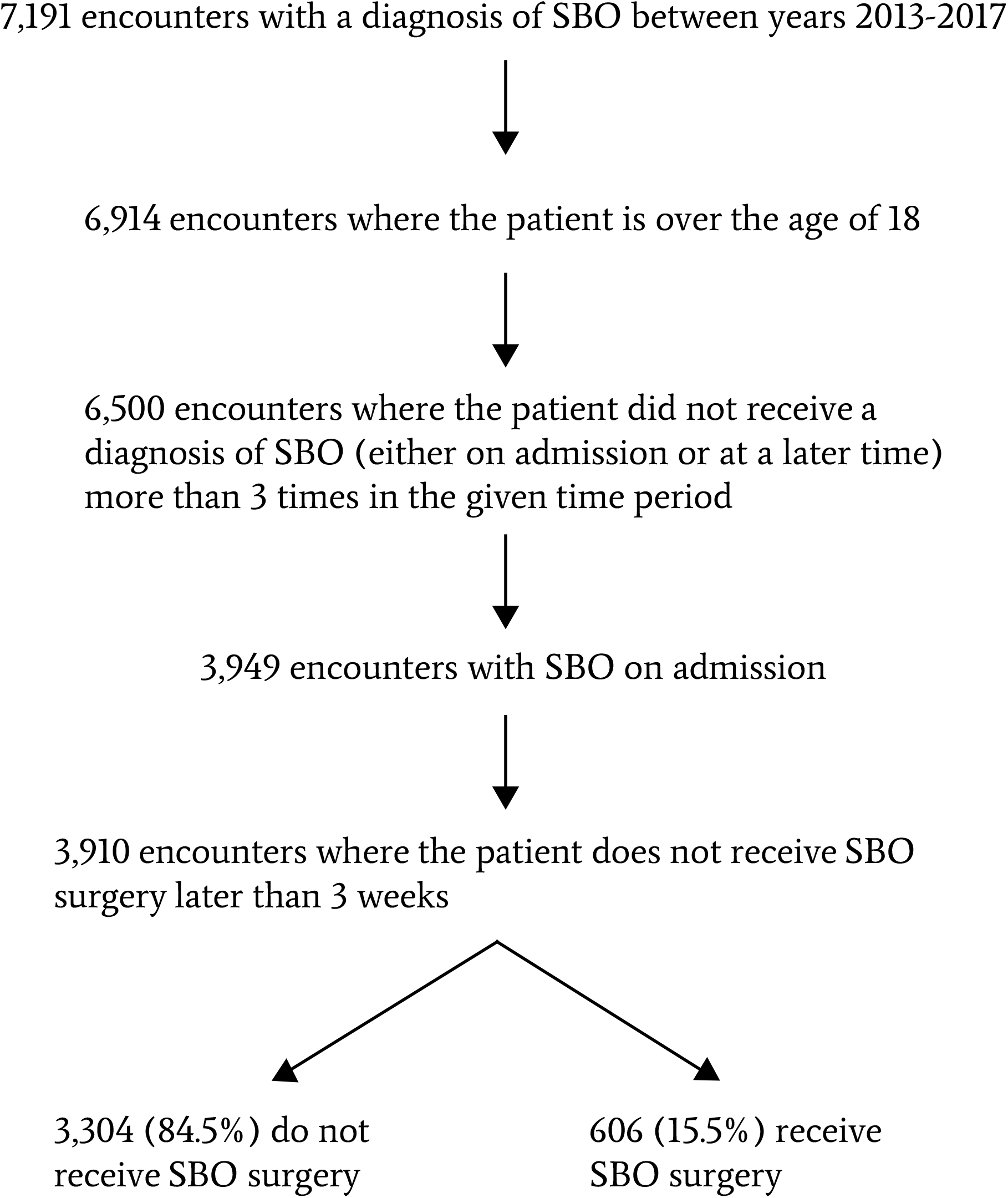
Stepwise Application of Exclusion Criteria.

**Figure 2:**
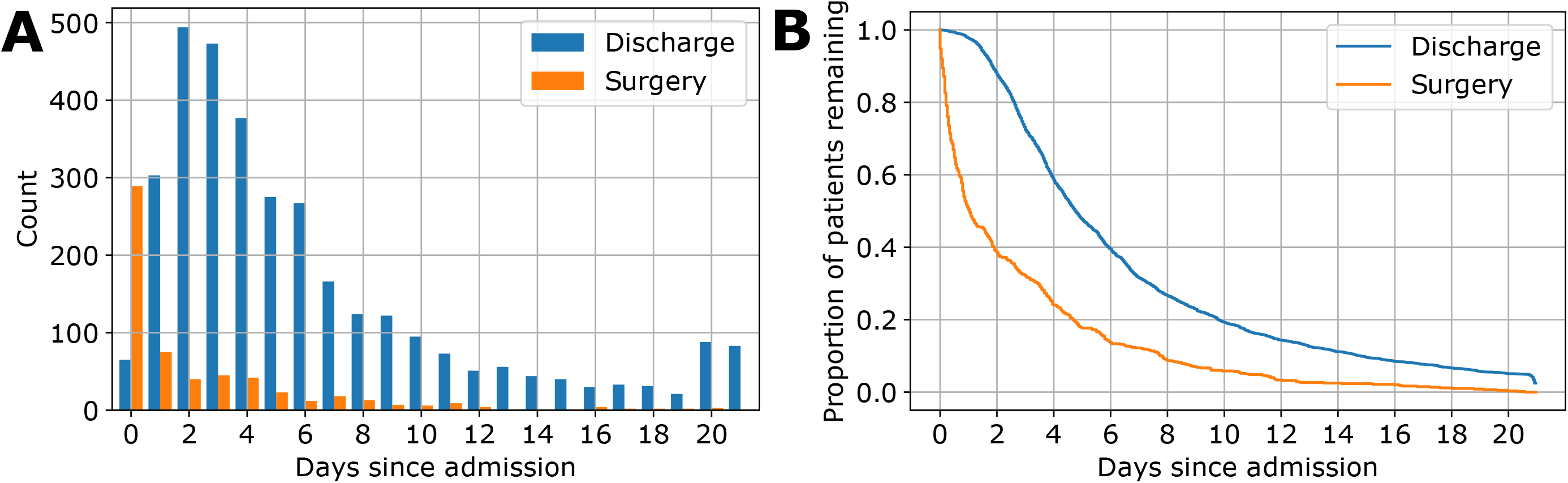
Encounter Endpoints. Panel (A) shows histograms of the time between admission and SBO-related surgery in the ‘Surgery’ group compared to the time between admission and discharge in the ‘Discharge’ group. Panel (B) shows the proportion of each group who have not yet received surgery or been discharged as a function of the time since admission.

### Description of Data and Missing Values

Each vital and laboratory measurement (Supplementary Table 1) was observed in over 80% of encounters (Supplementary Table 2). CT studies were conducted in 69.8% of all encounters (mean=1.16/encounter). Abdominal radiographs were taken in 62.0% of all encounters (mean=2.37/encounter). Small bowel follow through studies were conducted in 6.6% of all encounters (mean=1.02/encounter).

### Model Performance

Mean AUROC for the 24-hour prediction task increased from 0.644 at 12 hours after admission to 0.779 at 72 hours, with performance steadily improving between those times (see Figure 3, panels A-B). Similarly, performance for the within-encounter prediction task increased from 0.622 at 12 hours to 0.708 at 72 hours (see Figure 3, panels C-D), and the C-Index increased from 0.639 at 12 hours to 0.729 at 72 hours (Supplementary Figure 1). Peak performance for the 24-hour task (0.824) was observed at 108 hours after admission, whereas peak performance for the within-encounter task (0.712) and C-Index (0.735) were observed at 96 hours after admission, respectively. Model performance is shown in Figure 3. AUROC and C-Index values for all folds are presented in Supplementary Tables 4-8. See Supplementary Figure 2 for prediction performance with respect to time to event. Model coefficients are provided in Supplementary Table 9.

**Figure 3:**
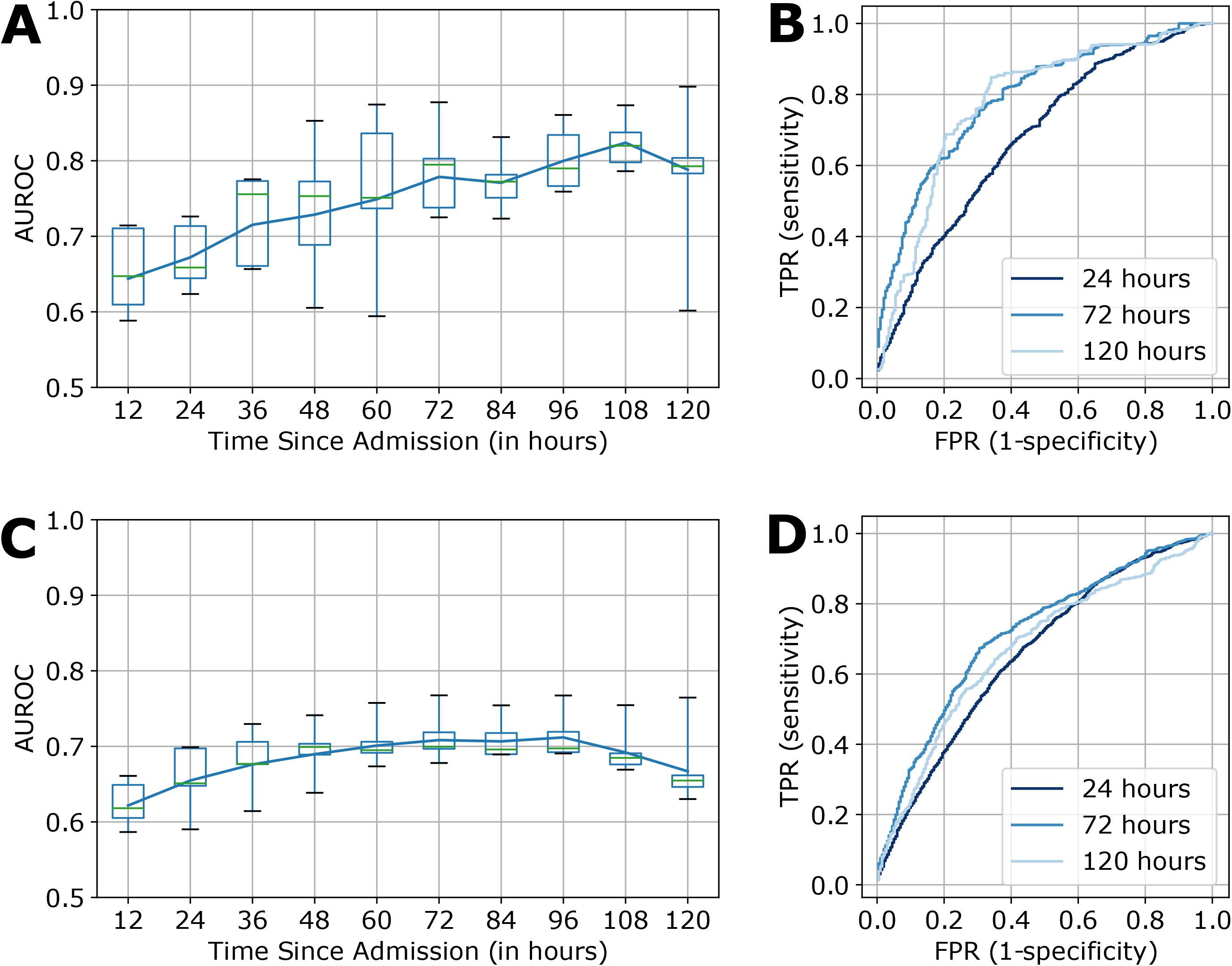
Prediction Performance over Time. Performance over time is quantified when predicting whether patients will receive surgery in the next 24 hours (*i*.*e*., “24-hour prediction”) and at any time during the current encounter (*i*.*e*., “within-encounter prediction”), respectively. The left panels show the area under the receiver operating characteristic curve (AUROC) by time since admission for (A) the 24-hour task, and (C) the within-encounter task. The line plot indicates the mean AUROC at each time point and the boxplots show values across all folds, with the green center lines indicating median values, box edges indicating the interquartile range, and whiskers indicating the maximum and minimum values. The right panels show the receiver operating characteristic (ROC) curve at 24, 72, and 120 hours since admission for (B) the 24-hour task, and (D) the within-encounter task.

High variance across folds for the 24-hour task can be explained by small numbers of positive classes in the test set in each time window as time progresses (see Figure 2). At 72 hours, for example, the number of surgeries occurring in the next 24 hours in each test fold ranges from three to six. For this reason, performance could not be reliably assessed beyond 120 hours after admission.

### Reduced Hospitalization Length

The observed incorrect discharge rate in our cohort was 9.3% (56 of 598 cases). When setting the discharge threshold to match this value, the model’s true discharge rate was 50.4%, the discharge predictive value was 96.7%, and the average reduction in hospital stay was approximately 63 hours (see Figure 4). When setting the threshold to achieve an incorrect discharge rate of 2.0%, the average reduction in hospital stay was approximately 18 hours.

**Figure 4:**
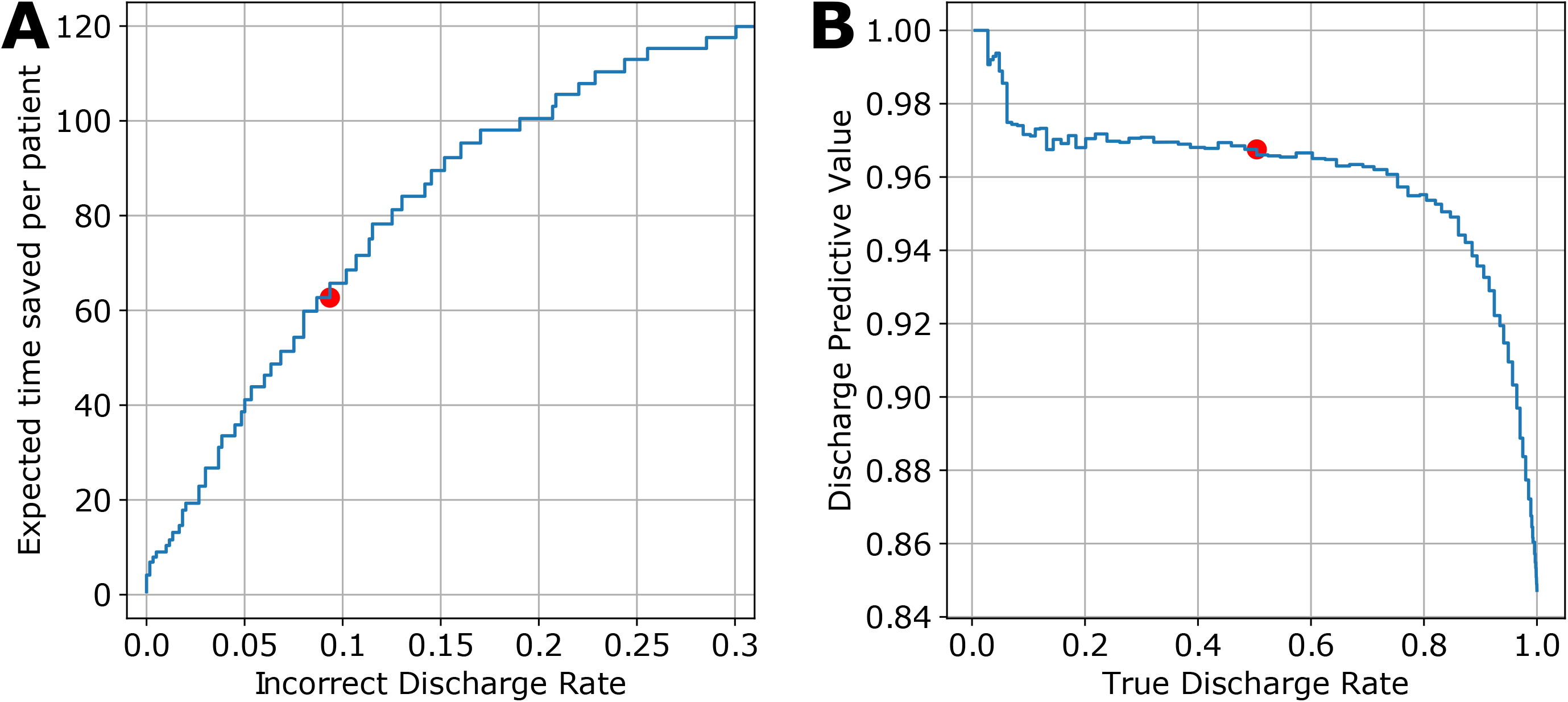
Discharge Prediction Performance. Panel (A) shows the average reduction in length of stay among patients eventually discharged versus the corresponding incorrect discharge rate. The red marker indicates the point on the curve that matches the incorrect discharge rate observed in our dataset as a consequence of clinician decision-making (9.4%). Panel (B) shows the tradeoff between discharge predictive value and the true discharge rate for the discharge prediction task. As before, the red marker indicates the point on the curve associated with clinician decision-making in the current cohort.

## DISCUSSION

This study is the first to develop a machine learning model for determining the risk of requiring surgery among patients admitted for SBO. The model was developed using a large dataset comprising 3,910 SBO-related encounters at three hospitals within the Duke University Health System over a five-year period. In contrast to earlier scoring systems, the proposed model integrates information from multiple clinical data sources (*e*.*g*., laboratory values, vital signs, inputs and outputs, radiological findings) to assign each patient a single score summarizing their relative risk. This risk score is time sensitive, changing as more clinical information is collected, to allow providers to dynamically assess patient status. Moreover, because the model determines relative risk rather than predicting surgery versus no surgery, it could be used to identify which cases are most urgent when prioritizing operating room resources, or to drive decisions about hospital resource allocation. It could also be used to determine which patients can safely be managed by a medical service versus a surgical service, or which patients should be transferred to a higher level of care or a center with surgical expertise. Not surprisingly, model performance steadily improves as more data are collected. Peak performance on both the 24-hour and within-encounter prediction tasks was observed approximately three days after admission. Prediction performance is less robust in the first 24 hours, likely due to the limited amount of clinical data available at that time. Although many cases result in surgery within the first 24 hours, these cases are often immediately evaluated as urgent and therefore proceed to surgery before laboratory results, radiological findings, and other informative data have been collected. Due to the model’s reliance on these findings, its greatest value may be in identifying whether patients not immediately requiring surgery remain at risk of deteriorating over time. In contrast, patients identified as low-risk by the model are unlikely to require surgery, and may be considered for earlier discharge.

Model-based discharge results confirm that identifying low-risk patients may be the most clinically actionable application of this work. On average, the model recommended discharge over 60 hours before discharge took place while maintaining an incorrect discharge rate lower than the true rate of SBO-related readmission in our cohort. It is impossible to determine precisely when the decision to discharge took place, but our experience suggests that this decision precedes discharge itself by several hours, not days. Thus, we conservatively estimate that model-guided discharge could reduce the average length of stay by over 2 days among patients not requiring surgery. Given the large number of SBO-related admissions per year, this could substantially reduce hospital costs, reduce costs to the patient, and improve patient satisfaction for hundreds of thousands of admissions in the US. Moreover, our simple decision rule for early discharge – namely, applying the same risk threshold across all time points -- should be fine-tuned to further improve performance in a real clinical deployment.

The next phase of this research is to prospectively deploy the model in a clinical setting. The model will be applied to all admitted patients with SBOs, providing near-real-time identification of patients predicted to require surgical intervention versus those who can be safely medically managed. An initial period of silent deployment, in which model predictions are not presented to clinicians, will be used to compare predictions with clinician decisions and patient outcomes to allow the model to be further validated and refined. If successful, the silent deployment will be followed by a prospective trial to evaluate the potential benefit of providing model-based recommendations to clinical teams. Model predictions would be used to inform provider decision-making, not replace it, and instances in which the team disagreed with the model’s prediction would be logged and examined. Our long-term goal is to integrate this model into our electronic health record to be incorporated in the normal clinician workflow.

### Limitations

This study has several limitations related to the retrospective nature of the analysis. Although thoughtful extraction and data cleaning procedures were applied, there was no mechanism to verify that clinical measurements were accurate or complete. To improve the accuracy, the patient selection was based on retrospective billing data. Of course, to implement this model would require real-time identification of these patients. Future iterations will define parameters for the patient population for whom this model may be useful. This should be studied prospectively. Additionally, the model was developed using data from DUHS only, and may not generalize effectively to another hospital system. Prior to deployment at another location, the model should be prospectively tested and/or refined. It is also recognized that additional time spent in the hospital may influence the ultimate outcome of a discharge, and a more granular approach to prediction may be needed to guide overall health support (intravenous fluid, *etc*.) in order to optimize outcomes. CT scans are integral to the management of SBOs, but we opted to omit imaging for this iteration of the model. This omission was felt to be justifiable as the aim of this study was to augment clinical decision-making, not to replicate prior studies about the role of imaging in determining which SBOs are operative. The ideal model would complement existing management practices by providing insights into trends in physiological measurements and how they relate to surgery risk, not replace more holistic management practices. Finally, this work assumes that patients deemed appropriate for surgical management were in fact appropriate for surgical management. We believe this is a reasonable assumption, however, our model has undoubtedly captured DUHS-specific clinical management strategies that may limit its generalizability. Further investigation is required, and a prospective randomized trial would be the optimal method to accomplish this.

## CONCLUSIONS

This work demonstrates that a machine learning approach can be used to continuously, effectively stratify patients in a large health system by their risk of requiring SBO surgery. The model effectively determined whether patients would require surgery within the next 24 hours and within the current encounter. Performance improved as more data was collected, peaking approximately 3 days after admission. Given this trend in model performance and the high number of surgeries occurring within 24 hours of admission, the model is most promising as a means to identify low-risk patients for earlier discharge. Model predictions can also be used to prioritize operating room and hospital resources, or to determine which patients should be managed by medical versus surgical services or transferred to a higher level of care. Future study will quantify model performance when applied to other hospital systems and explore the benefit of a prospective deployment in an inpatient setting.

## Data Availability

Data not available due to privacy restrictions.

## ADDITIONAL INFORMATION

### Author Contributions

Study conception and design: JW, DT, AK

Acquisition of data: JW, DT

Analysis of data: MT

Interpretation of data: MT, ME, RH, LC

Drafting of manuscript: MT, JW, ME

Critical revision: LC, AK, RH

## REFERENCES

1. Aquina, C.T., Becerra, A.Z., Probst, C.P., et al. Patients with adhesive small bowel obstruction should be primarily managed by a surgical team: Ann Surg. 2016;264(3):437–447. doi:10.1097/SLA.0000000000001861

2. Scott, J.W., Olufajo, O.A., Brat, G.A., et al. Use of national burden to define operative emergency general surgery. JAMA Surg. 2016;151(6):e160480. doi:10.1001/jamasurg.2016.0480

3. Duron, J.-J., Silva, N.J.-D., du Montcel, S.T., et al. Adhesive postoperative small bowel obstruction: incidence and risk factors of recurrence after surgical treatment: a multicenter prospective study. Ann Surg. 2006;244(5):750–757. doi:10.1097/01.sla.0000225097.60142.68

4. Miller, G., Boman, J., Shrier, I., Gordon, P.H. Etiology of small bowel obstruction. Am J Surg. 2000;180(1):33–36. doi:10.1016/S0002-9610(00)00407-4

5. Foster, N.M., McGory, M.L., Zingmond, D.S., Ko, C.Y. Small bowel bbstruction: a population-based appraisal. J Am Coll Surg. 2006;203(2):170–176. doi:10.1016/j.jamcollsurg.2006.04.020

6. Fevang, B.T., Fevang, J.M., Søreide, O., Svanes, K., Viste, A. Delay in operative treatment among patients with small bowel obstruction. Scand J Surg. 2003;92(2):131–137. doi:10.1177/145749690309200204

7. Keenan, J.E., Turley, R.S., McCoy, C.C., Migaly, J., Shapiro, M.L., Scarborough, J.E. Trials of nonoperative management exceeding 3 days are associated with increased morbidity in patients undergoing surgery for uncomplicated adhesive small bowel obstruction. J Trauma Acute Care Surg. 2014;76(6):1367–1372. doi:10.1097/TA.0000000000000246

8. Schraufnagel, D., Rajaee, S., Millham, F.H. How many sunsets? Timing of surgery in adhesive small bowel obstruction: a study of the nationwide inpatient sample. J Trauma Acute Care Surg. 2013;74(1):181–189. doi:10.1097/TA.0b013e31827891a1

9. Bilderback, P.A., Massman, J.D., Smith, R.K., La Selva, D., Helton, W.S. Small bowel obstruction is a surgical disease: patients with adhesive small bowel obstruction requiring operation have more cost-effective care when admitted to a surgical service. J Am Coll Surg. 2015;221(1):7–13. doi:10.1016/j.jamcollsurg.2015.03.054

10. Malangoni, M.A., Times, M.L., Kozik, D., Merlino, JI. Admitting service influences the outcomes of patients with small bowel obstruction. Surgery. 2001;130(4):706–713. doi:10.1067/msy.2001.116918

11. Cox, D.R. Regression models and life-tables. J R Stat Soc Ser B Methodol. 1972;34(2):187–220.

12. Balakumar, B.J., Hastie, T., Friedman, J., Tibshirani Simon, N.R. Glmnet for Python.; 2016. http://www.stanford.edu/~hastie/glmnet_python/.

13. Varma, S., Simon, R. Bias in error estimation when using cross-validation for model selection. BMC Bioinformatics. 2006;7(1):91. doi:10.1186/1471-2105-7-91

14. Bradley, A.P. The use of the area under the ROC curve in the evaluation of machine learning algorithms. Pattern Recognition. Pattern Recognition. 1997;30:1145–1159. doi:10.1016/S0031-3203(96)00142-2

15. Harrell, F.E., Lee, K.L., Mark, D.B. Multivariable prognostic models: issues in developing models, evaluating assumptions and adequacy, and measuring and reducing errors. Stat Med. 1996;15(4):361–387. doi:10.1002/(SICI)1097-0258(19960229)15:4<361::AID-SIM168>3.0.CO;2-4

16. Arung, W. Pathophysiology and prevention of postoperative peritoneal adhesions. World J Gastroenterol. 2011;17(41):4545. doi:10.3748/wjg.v17.i41.4545

