## Supplementary Tables and Figures for "Machine Learning Prediction of Surgical Intervention for Small Bowel Obstruction"

**Supplementary Table 1:** Overview of variables

**Supplementary Table 2:** Variable Descriptive Statistics and Observation Rates

**Supplementary Figure 1:** Concordance Index over Time

**Supplementary Figure 2:** Prediction Performance Relative to Time to Event

**Supplementary Table 3:** Words and phrases selected by expert panel

**Supplementary Table 4:** AUROC over Time for the 24-hour Prediction Task

**Supplementary Table 5:** AUROC over Time for Surgery at any Point

**Supplementary Table 6:** Concordance Index over Time

**Supplementary Table 7:** AUROC Relative to Time to Event

**Supplementary Table 8:** Concordance Index Relative to Time to Event

**Supplementary Table 9**: Model Coefficients

**Supplementary Table 1: Overview of variables**

| Category | Variable | Description |
| --- | --- | --- |
|  | Age | Age at time of encounter |
|  | Location | Duke University, Duke Regional, Duke Raleigh |
|  | Number of Past SBOs |  |
|  | Hour Since Admission | Based on Admission Timestamp |
|  | Time of Day |  |
| Vitals | Diastolic Blood Pressure | Vital Sign Collected on Admission |
|  | Systolic Blood Pressure | Vital Sign Collected on Admission |
|  | Pulse | Vital Sign Collected on Admission |
|  | Respiratory Rate | Vital Sign Collected on Admission |
|  | SPO2 | Vital Sign Collected on Admission |
| Labs | Albumin | Serum Level from Metabolic Panel |
|  | Calcium | Serum Level |
|  | Carbon Dioxide | Serum Level |
|  | Chloride | Serum Level from Metabolic Panel |
|  | Creatinine | Serum Level from Metabolic Panel |
|  | Hematocrit | Serum Level |
|  | Hemoglobin | Serum Level from CBC |
|  | Platelet Count | Serum Level from CBC |
|  | Potassium | Serum Level from Metabolic Panel |
|  | Sodium | Serum Level from Metabolic Panel |
|  | White Blood Cell Count | Serum Level from CBC |
| Inputs/outputs | Maintenance IV Volume | Intravenous Fluid Intake |
|  | Piggyback IV Volume | Intravenous Fluid Intake |
|  | Tube Intake | Nasogastric Tube Intake |
|  | Tube Output | Nasogastric Tube Output |
|  | Urine Output | Urine Output w/ and w/o catheter |
|  | Emesis Occurrence | Emesis Present or not |
|  | Stool Occurrence | Stool Output or not |
|  | Urine Occurrence | Urine Output or not |

**Supplementary Table 2: Variable Descriptive Statistics and Observation Rates**

| Variable | Mean ± SD (range) | Encounters with at least one measurement, N (%) |
| --- | --- | --- |
| Diastolic Blood Pressure (mmHg) | 70.2±13.6 (0-171) | 3901 (99.8%) |
| Systolic Blood Pressure (mmHg) | 127.8±23.4 (0-476) | 3901 (99.8%) |
| Pulse (bpm) | 87.9±18.5 (0-369) | 3906 (99.9%) |
| Respiratory Rate (rpm) | 19.2±4.8 (0-189) | 3905 (99.9%) |
| SPO2 (%) | 96.6±3.0 (0-100) | 3904 (99.8%) |
| Platelet Count (K/mcL) | 250.4±136.8 (1-2442) | 3507 (89.7%) |
| White Blood Cell Count (K/mcL) | 10.1±6.6 (0.1-117.6) | 3508 (89.7%) |
| Hemoglobin (g/dL) | 10.5±2.1 (2.9-20.3) | 3508 (89.7%) |
| Hematocrit (%) | 15.5±16.3 (9-58.9) | 3188 (81.5%) |
| Sodium (mmol/L) | 137.7±4.9 (110-177) | 3818 (97.6%) |
| Potassium (mmol/L) | 3.9±0.6 (1.5-9.1) | 3814 (97.5%) |
| Chloride (mmol/L) | 104.4±6.5 (54-155) | 3818 (97.6%) |
| Albumin (g/dL) | 2.9±0.9 (1-6) | 3310 (84.7%) |
| Calcium (mg/dL) | 8.5±0.7 (3.4-14.4) | 3818 (97.6%) |
| Carbon Dioxide (mmol/L) | 25.3±4.2 (5-50) | 3818 (97.6%) |
| Creatinine (mg/dL) | 1.4±1.4 (0.1-26.8) | 3818 (97.6%) |

**Supplementary Figure 1: Concordance Index over Time.** Boxplots depict the concordance index (c-index) across all folds by time since admission, with the green center lines indicating median values, box edges indicating the interquartile range, and whiskers indicating the maximum and minimum values. The line plot indicates the mean c-index at each time point.

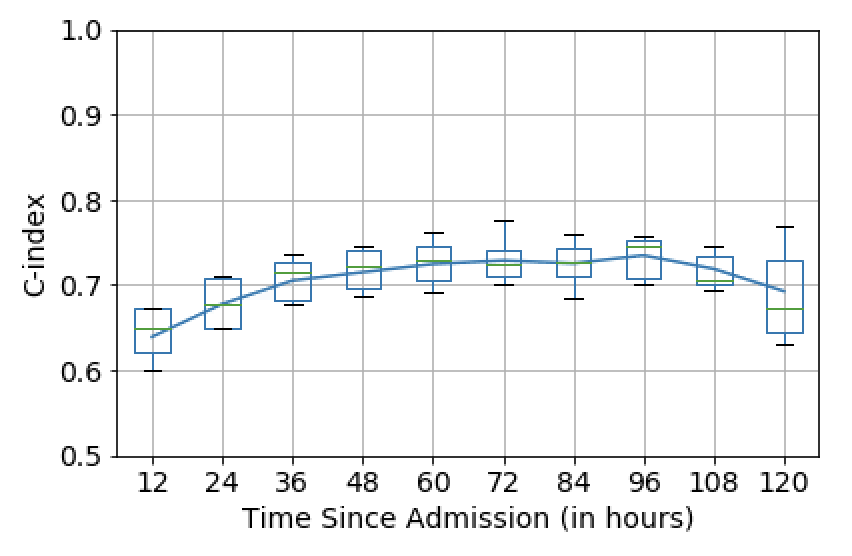

**Supplementary Figure 2: Prediction Performance Relative to Time to Event.** Panels (A, B) show model performance by time until surgery or discharge when predicting whether the patient will receive surgery at any time during the current encounter. In panel (A), performance is quantified as the area under the receiver operating characteristic curve (AUROC), while in panel (B), performance is quantified as the concordance index (c-index). Boxplots depict model performance values across all folds, with the green center lines indicating median values, box edges indicating the interquartile range, and whiskers indicating the maximum and minimum values. The line plot indicates the mean performance value at each time point.

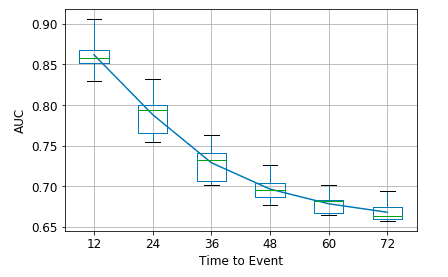

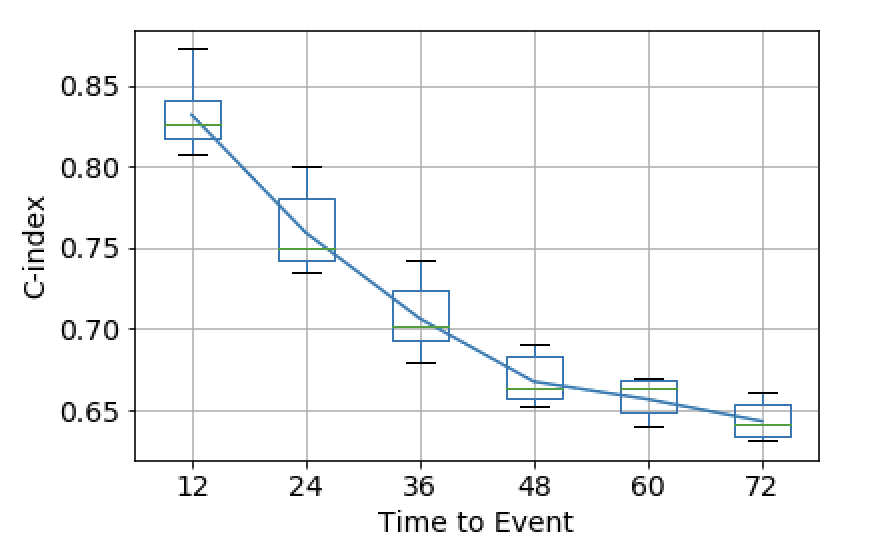

**Supplementary Table 3: Words and phrases identified as clinically meaningful and interpretable.** The table includes common words and phrases from CT and small bowel follow through studies, respectively, and whether they are more strongly associated with SBO surgery (‘Surgery’) or no SBO surgery (‘Discharge’) in our dataset.

| **CT** | | **Small bowel follow through** | |
| --- | --- | --- | --- |
| **Surgery** | **Discharge** | **Surgery** | **Discharge** |
| ischemia | ileus |  | partial |
| Closed loop | Early or partial |  | Normal transit |
| Internal hernia | No ascites |  | Bowel transit |
| Transition point | unchanged |  | Gaseous distention |
| Pneumotosis or free |  |  | Normal appearance |
| Impression high grade |  |  | Opacification of the |

**Supplementary Table 4: AUROC over Time for the 24-hour Prediction Task.** Values for all folds presented in Figure 3, Panel (A) are provided in the table below.

|  | Time Since Admission (in hours) | | | | | | | | | |
| --- | --- | --- | --- | --- | --- | --- | --- | --- | --- | --- |
|  | 12 | 24 | 36 | 48 | 60 | 72 | 84 | 96 | 108 | 120 |
| Fold 1 | 0.65 | 0.71 | 0.76 | 0.77 | 0.84 | 0.80 | 0.75 | 0.79 | 0.87 | 0.78 |
| Fold 2 | 0.61 | 0.62 | 0.66 | 0.69 | 0.75 | 0.88 | 0.83 | 0.86 | 0.84 | 0.60 |
| Fold 3 | 0.71 | 0.73 | 0.77 | 0.85 | 0.87 | 0.80 | 0.72 | 0.76 | 0.80 | 0.80 |
| Fold 4 | 0.71 | 0.66 | 0.78 | 0.75 | 0.74 | 0.74 | 0.78 | 0.83 | 0.82 | 0.90 |
| Fold 5 | 0.59 | 0.64 | 0.66 | 0.61 | 0.59 | 0.73 | 0.77 | 0.77 | 0.79 | 0.79 |
| Mean | 0.64 | 0.67 | 0.72 | 0.73 | 0.75 | 0.78 | 0.77 | 0.80 | 0.82 | 0.79 |
| SD | 0.06 | 0.04 | 0.06 | 0.09 | 0.11 | 0.06 | 0.04 | 0.04 | 0.03 | 0.11 |

**Supplementary Table 5: AUROC over Time for Surgery at any Point.** Values for all folds presented in Figure 3, Panel (B) are provided in the table below.

|  | Time Since Admission (in hours) | | | | | | | | | |
| --- | --- | --- | --- | --- | --- | --- | --- | --- | --- | --- |
|  | 12 | 24 | 36 | 48 | 60 | 72 | 84 | 96 | 108 | 120 |
| Fold 1 | 0.65 | 0.70 | 0.71 | 0.70 | 0.69 | 0.70 | 0.69 | 0.69 | 0.69 | 0.65 |
| Fold 2 | 0.59 | 0.65 | 0.68 | 0.70 | 0.71 | 0.72 | 0.72 | 0.72 | 0.69 | 0.66 |
| Fold 3 | 0.66 | 0.70 | 0.73 | 0.74 | 0.76 | 0.77 | 0.75 | 0.77 | 0.76 | 0.77 |
| Fold 4 | 0.62 | 0.59 | 0.61 | 0.64 | 0.67 | 0.68 | 0.69 | 0.69 | 0.67 | 0.66 |
| Fold 5 | 0.61 | 0.65 | 0.68 | 0.69 | 0.70 | 0.70 | 0.70 | 0.70 | 0.68 | 0.63 |
| Mean | 0.62 | 0.66 | 0.68 | 0.69 | 0.70 | 0.71 | 0.71 | 0.71 | 0.69 | 0.67 |
| SD | 0.03 | 0.04 | 0.04 | 0.04 | 0.03 | 0.03 | 0.03 | 0.03 | 0.03 | 0.05 |

**Supplementary Table 6: Concordance Index over Time**

|  | Time Since Admission (in hours) | | | | | | | | | |
| --- | --- | --- | --- | --- | --- | --- | --- | --- | --- | --- |
|  | 12 | 24 | 36 | 48 | 60 | 72 | 84 | 96 | 108 | 120 |
| Fold 1 | 0.65 | 0.71 | 0.73 | 0.72 | 0.71 | 0.70 | 0.69 | 0.70 | 0.70 | 0.63 |
| Fold 2 | 0.62 | 0.68 | 0.72 | 0.75 | 0.76 | 0.78 | 0.76 | 0.75 | 0.69 | 0.64 |
| Fold 3 | 0.67 | 0.71 | 0.74 | 0.74 | 0.75 | 0.74 | 0.73 | 0.75 | 0.75 | 0.77 |
| Fold 4 | 0.67 | 0.65 | 0.68 | 0.70 | 0.73 | 0.72 | 0.74 | 0.76 | 0.73 | 0.73 |
| Fold 5 | 0.60 | 0.65 | 0.68 | 0.69 | 0.69 | 0.71 | 0.71 | 0.71 | 0.71 | 0.67 |
| Mean | 0.64 | 0.68 | 0.71 | 0.72 | 0.73 | 0.73 | 0.73 | 0.74 | 0.72 | 0.69 |
| SD | 0.03 | 0.03 | 0.03 | 0.03 | 0.03 | 0.03 | 0.03 | 0.03 | 0.02 | 0.06 |

**Supplementary Table 7: AUROC Relative to Time to Event.** Values for all folds presented in Supplementary Figure 2, Panel (A) are provided in the table below.

|  | Time to Event (in hours) | | | | | |
| --- | --- | --- | --- | --- | --- | --- |
|  | 12 | 24 | 36 | 48 | 60 | 72 |
| Fold 1 | 0.87 | 0.81 | 0.73 | 0.70 | 0.68 | 0.67 |
| Fold 2 | 0.85 | 0.78 | 0.72 | 0.67 | 0.65 | 0.65 |
| Fold 3 | 0.91 | 0.83 | 0.75 | 0.71 | 0.68 | 0.67 |
| Fold 4 | 0.85 | 0.76 | 0.70 | 0.67 | 0.66 | 0.65 |
| Fold 5 | 0.84 | 0.76 | 0.71 | 0.68 | 0.67 | 0.65 |
| Mean | 0.86 | 0.79 | 0.72 | 0.69 | 0.67 | 0.65 |
| SD | 0.03 | 0.03 | 0.02 | 0.02 | 0.01 | 0.01 |

**Supplementary Table 8: Concordance Index Relative to Time to Event**

|  | Time to Event (in hours) | | | | | |
| --- | --- | --- | --- | --- | --- | --- |
|  | 12 | 24 | 36 | 48 | 60 | 72 |
| Fold 1 | 0.84 | 0.78 | 0.72 | 0.68 | 0.67 | 0.66 |
| Fold 2 | 0.82 | 0.75 | 0.70 | 0.66 | 0.64 | 0.63 |
| Fold 3 | 0.87 | 0.80 | 0.74 | 0.69 | 0.67 | 0.65 |
| Fold 4 | 0.83 | 0.74 | 0.68 | 0.65 | 0.65 | 0.64 |
| Fold 5 | 0.81 | 0.74 | 0.69 | 0.66 | 0.66 | 0.63 |
| Mean | 0.83 | 0.76 | 0.71 | 0.67 | 0.66 | 0.64 |
| SD | 0.03 | 0.03 | 0.03 | 0.02 | 0.01 | 0.01 |

**Supplementary Table 9: Model Coefficients.** Statistics for the distribution of the model coefficients over the 5 cross validation folds are provided below. For exponential moving averages/sums, the value in window corresponds to the half-life. For indicator variables, the value in window indicates how many hours a positive value would be carried forward before returning to zero.

| Group | Variable | Calculated Value | Window | Mean | Std | Min | Max |
| --- | --- | --- | --- | --- | --- | --- | --- |
| Encounter | Age | Fixed |  | 0.0193 | 0.0083 | 0.0079 | 0.0284 |
|  | Duke Loc | Fixed |  | -0.0271 | 0.0124 | -0.0432 | 0.0124 |
|  | Hour since admission | Current value |  | -0.0516 | 0.0239 | -0.0922 | 0.0239 |
|  | Past SBO | Fixed |  | 0.0232 | 0.0255 | -0.0266 | 0.0459 |
|  | Raleigh Loc | Fixed |  | 0.0919 | 0.0142 | 0.0142 | 0.1127 |
|  | Regional Loc | Fixed |  | -0.0311 | 0.0146 | -0.051 | 0.0146 |
|  | Time of Day | Current value |  | -0.0001 | 0.0001 | -0.0003 | 0.0001 |
| Vitals | BP Diastolic | Current value |  | 0.024 | 0.0054 | 0.0054 | 0.0337 |
|  |  | Exp moving avg | 24 | 0.0002 | 0.0009 | -0.0015 | 0.0009 |
|  |  |  | 6 | -0.0026 | 0.0003 | -0.0028 | 0.0003 |
|  |  |  | 72 | 0.0017 | 0.0015 | -0.0009 | 0.0029 |
|  |  | Time since last measurement |  | 0.0099 | 0.0149 | -0.0197 | 0.0199 |
|  | BP Systolic | Current value |  | 0.0185 | 0.0034 | 0.0034 | 0.0243 |
|  |  | Exp moving avg | 24 | 0.0024 | 0.0011 | 0.0005 | 0.0039 |
|  |  |  | 6 | -0.0006 | 0.0006 | -0.0012 | 0.0006 |
|  |  |  | 72 | 0.0037 | 0.0014 | 0.0011 | 0.0052 |
|  |  | Time since last measurement |  | 0.0099 | 0.0149 | -0.0197 | 0.0199 |
|  | Pulse | Current value |  | 0.0218 | 0.0041 | 0.0041 | 0.0271 |
|  |  | Exp moving avg | 24 | 0.0106 | 0.0016 | 0.0016 | 0.0124 |
|  |  |  | 6 | 0.001 | 0.0004 | 0.0004 | 0.0015 |
|  |  |  | 72 | 0.0172 | 0.0032 | 0.0032 | 0.0209 |
|  |  | Time since last measurement |  | -0.0064 | 0.0063 | -0.0154 | 0.0063 |
|  | Respiratory Rate | Current value |  | -0.0022 | 0.0055 | -0.0086 | 0.0075 |
|  |  | Exp moving avg | 24 | 0.0027 | 0.0006 | 0.0006 | 0.0033 |
|  |  |  | 6 | -0.0009 | 0.001 | -0.0018 | 0.0011 |
|  |  |  | 72 | 0.0054 | 0.0016 | 0.0016 | 0.0073 |
|  |  | Time since last measurement |  | -0.0101 | 0.0066 | -0.0149 | 0.0066 |
|  | SPO2 | Current value |  | -0.0033 | 0.0042 | -0.0101 | 0.0042 |
|  |  | Exp moving avg | 24 | 0.0007 | 0.0006 | -0.0004 | 0.0014 |
|  |  |  | 6 | 0.001 | 0.0005 | 0.0001 | 0.0016 |
|  |  |  | 72 | 0.0012 | 0.0004 | 0.0004 | 0.0019 |
|  |  | Time since last measurement |  | 0.0012 | 0.0004 | 0.0004 | 0.0019 |
| Labs | Albumin | Current value |  | 0.0368 | 0.0043 | 0.0043 | 0.0415 |
|  |  | Exp moving avg | 12 | -0.0004 | 0.0007 | -0.0014 | 0.0007 |
|  |  |  | 144 | 0.0025 | 0.0008 | 0.0008 | 0.0038 |
|  |  |  | 48 | 0.0014 | 0.0008 | 0.0002 | 0.0023 |
|  |  | Time since last measurement |  | -0.0364 | 0.007 | -0.0449 | 0.007 |
|  | Calcium | Current value |  | 0.0234 | 0.0057 | 0.0057 | 0.0327 |
|  |  | Exp moving avg | 12 | -0.0028 | 0.0008 | -0.0043 | 0.0008 |
|  |  |  | 144 | -0.0006 | 0.0003 | -0.0011 | 0.0003 |
|  |  |  | 48 | -0.0013 | 0.0004 | -0.0021 | 0.0004 |
|  |  | Time since last measurement |  | -0.0106 | 0.0007 | -0.0111 | 0.0007 |
|  | Carbon Dioxide | Current value |  | 0.0039 | 0.0073 | -0.0081 | 0.0108 |
|  |  | Exp moving avg | 12 | 0.0008 | 0.0009 | -0.0006 | 0.0021 |
|  |  |  | 144 | -0.0002 | 0.0009 | -0.0019 | 0.0009 |
|  |  |  | 48 | 0.0002 | 0.0007 | -0.001 | 0.0011 |
|  |  | Time since last measurement |  | -0.0105 | 0.0006 | -0.011 | 0.0006 |
|  | Chloride | Current value |  | -0.006 | 0.007 | -0.0152 | 0.007 |
|  |  | Exp moving avg | 12 | -0.0005 | 0.0003 | -0.001 | 0.0003 |
|  |  |  | 144 | 0.0017 | 0.0008 | 0.0007 | 0.0026 |
|  |  |  | 48 | 0.0007 | 0.0006 | -0.0002 | 0.0015 |
|  |  | Time since last measurement |  | -0.0105 | 0.0006 | -0.011 | 0.0006 |
|  | Creatinine | Current value |  | -0.0208 | 0.01 | -0.0364 | 0.01 |
|  |  | Exp moving avg | 12 | -0.0002 | 0.0005 | -0.0008 | 0.0007 |
|  |  |  | 144 | -0.004 | 0.0015 | -0.0064 | 0.0015 |
|  |  |  | 48 | -0.0023 | 0.0008 | -0.0035 | 0.0008 |
|  |  | Time since last measurement |  | -0.0105 | 0.0006 | -0.011 | 0.0006 |
|  | Hematocrit | Current value |  | 0.0283 | 0.006 | 0.006 | 0.0386 |
|  |  | Exp moving avg | 12 | 0.002 | 0.0003 | 0.0003 | 0.0024 |
|  |  |  | 144 | 0.0039 | 0.0007 | 0.0007 | 0.005 |
|  |  |  | 48 | 0.0042 | 0.0004 | 0.0004 | 0.0047 |
|  |  | Time since last measurement |  | 0.0081 | 0.0062 | -0.0001 | 0.0167 |
|  | Hemoglobin | Current value |  | 0.0281 | 0.0042 | 0.0042 | 0.0357 |
|  |  | Exp moving avg | 12 | 0.0005 | 0.0003 | 0.0003 | 0.001 |
|  |  |  | 144 | 0.0017 | 0.0008 | 0 | 0.0023 |
|  |  |  | 48 | 0.0021 | 0.0005 | 0.0005 | 0.0025 |
|  |  | Time since last measurement |  | -0.0165 | 0.0047 | -0.0228 | 0.0047 |
|  | Platelet Count | Current value |  | -0.006 | 0.0091 | -0.0171 | 0.0091 |
|  |  | Exp moving avg | 12 | -0.0004 | 0.0006 | -0.0013 | 0.0006 |
|  |  |  | 144 | -0.0015 | 0.0003 | -0.0018 | 0.0003 |
|  |  |  | 48 | -0.0011 | 0.0001 | -0.0014 | 0.0001 |
|  |  | Time since last measurement |  | -0.0168 | 0.0047 | -0.0231 | 0.0047 |
|  | Potassium | Current value |  | 0.0047 | 0.0031 | -0.0005 | 0.0092 |
|  |  | Exp moving avg | 12 | 0.001 | 0.0009 | -0.0001 | 0.0023 |
|  |  |  | 144 | 0.004 | 0.001 | 0.001 | 0.0052 |
|  |  |  | 48 | 0.003 | 0.0009 | 0.0009 | 0.0042 |
|  |  | Time since last measurement |  | -0.0108 | 0.0006 | -0.0113 | 0.0006 |
|  | Sodium | Current value |  | 0.0069 | 0.0064 | -0.0042 | 0.0147 |
|  |  | Exp moving avg | 12 | -0.0013 | 0.0005 | -0.0019 | 0.0005 |
|  |  |  | 144 | 0.0007 | 0.0006 | -0.0004 | 0.0014 |
|  |  |  | 48 | 0.0002 | 0.0004 | -0.0005 | 0.0008 |
|  |  | Time since last measurement |  | -0.0107 | 0.0006 | -0.0112 | 0.0006 |
|  | White Blood Cell Count | Current value |  | -0.0057 | 0.0049 | -0.0126 | 0.0049 |
|  |  | Exp moving avg | 12 | 0.0022 | 0.0005 | 0.0005 | 0.0026 |
|  |  |  | 144 | 0.004 | 0.001 | 0.001 | 0.0057 |
|  |  |  | 48 | 0.0031 | 0.0008 | 0.0008 | 0.0044 |
|  |  | Time since last measurement |  | -0.0167 | 0.0047 | -0.023 | 0.0047 |
| I/O | Maintenance IV Volume | Current value |  | 0.0768 | 0.006 | 0.006 | 0.0825 |
|  |  | Exp moving sum | 24 | 0.0381 | 0.0044 | 0.0044 | 0.0451 |
|  |  |  | 6 | 0.0343 | 0.0024 | 0.0024 | 0.0381 |
|  |  |  | 72 | 0.0225 | 0.0113 | 0.0113 | 0.0435 |
|  |  | Time since last measurement |  | -0.0727 | 0.0043 | -0.0782 | 0.0043 |
|  | Piggyback IV Volume | Current value |  | -0.0056 | 0.0082 | -0.0214 | 0.0082 |
|  |  | Exp moving sum | 24 | -0.0131 | 0.0099 | -0.0284 | 0.0099 |
|  |  |  | 6 | 0.0004 | 0.005 | -0.0088 | 0.005 |
|  |  |  | 72 | -0.0231 | 0.0117 | -0.0427 | 0.0117 |
|  |  | Time since last measurement |  | -0.0178 | 0.0044 | -0.0251 | 0.0044 |
|  | Tube Intake | Current value |  | -0.1072 | 0.0179 | -0.1313 | 0.0179 |
|  |  | Exp moving sum | 24 | -0.0048 | 0.0018 | -0.0072 | 0.0018 |
|  |  |  | 6 | -0.0063 | 0.0027 | -0.0106 | 0.0027 |
|  |  |  | 72 | -0.0042 | 0.0015 | -0.0054 | 0.0015 |
|  |  | Time since last measurement |  | -0.0065 | 0.0057 | -0.0161 | 0.0057 |
|  | Tube Output | Current value |  | 0.1739 | 0.0053 | 0.0053 | 0.182 |
|  |  | Exp moving sum | 24 | 0.0815 | 0.0041 | 0.0041 | 0.0879 |
|  |  |  | 6 | 0.0293 | 0.0007 | 0.0007 | 0.0304 |
|  |  |  | 72 | 0.0872 | 0.0068 | 0.0068 | 0.0965 |
|  |  | Time since last measurement |  | -0.0887 | 0.0054 | -0.0949 | 0.0054 |
|  | Urine Output | Current value |  | 0.0114 | 0.0235 | -0.0332 | 0.0348 |
|  |  | Exp moving sum | 24 | -0.0175 | 0.0115 | -0.0364 | 0.0115 |
|  |  |  | 6 | -0.0078 | 0.0071 | -0.0199 | 0.0071 |
|  |  |  | 72 | -0.0206 | 0.0144 | -0.0463 | 0.0144 |
|  |  | Time since last measurement |  | -0.0109 | 0.0112 | -0.0298 | 0.0112 |
| I/O Occurrence | Emesis | Current value |  | 0.0517 | 0.0039 | 0.0039 | 0.058 |
|  |  | Exp moving sum | 24 | 0.1041 | 0.0115 | 0.0115 | 0.1241 |
|  |  |  | 6 | 0.0994 | 0.0146 | 0.0146 | 0.1271 |
|  |  |  | 72 | 0.0798 | 0.0126 | 0.0126 | 0.0968 |
|  |  | Time since last measurement |  | -0.0366 | 0.0058 | -0.0456 | 0.0058 |
|  | Stool | Current value |  | -0.053 | 0.0045 | -0.0584 | 0.0045 |
|  |  | Exp moving sum | 24 | -0.0252 | 0.0018 | -0.0274 | 0.0018 |
|  |  |  | 6 | -0.1005 | 0.0058 | -0.1091 | 0.0058 |
|  |  |  | 72 | -0.0593 | 0.0026 | -0.0626 | 0.0026 |
|  |  | Time since last measurement |  | 0.0125 | 0.0042 | 0.0042 | 0.0173 |
|  | Urine | Current value |  | -0.0238 | 0.006 | -0.0315 | 0.006 |
|  |  | Exp moving sum | 24 | -0.0104 | 0.0024 | -0.0141 | 0.0024 |
|  |  |  | 6 | -0.0442 | 0.0097 | -0.059 | 0.0097 |
|  |  |  | 72 | -0.0208 | 0.0054 | -0.0296 | 0.0054 |
|  |  | Time since last measurement |  | -0.0168 | 0.0058 | -0.0251 | 0.0058 |
| SBO Follow Through | Imaging recorded | Indicator | 24 | 0.1976 | 0.0409 | 0.0409 | 0.2659 |
|  |  |  | 504 | 0.074 | 0.0323 | 0.0323 | 0.1265 |
|  | Bowel Transit | Indicator | 24 | 0 | 0 | 0 | 0 |
|  |  |  | 504 | 0 | 0 | 0 | 0 |
|  | Gaseous Distention | Indicator | 24 | 0 | 0 | 0 | 0 |
|  |  |  | 504 | 0 | 0 | 0 | 0 |
|  | Normal Appearance | Indicator | 24 | 0 | 0 | 0 | 0 |
|  |  |  | 504 | 0 | 0 | 0 | 0 |
|  | Normal Transit | Indicator | 24 | 0 | 0 | 0 | 0 |
|  |  |  | 504 | 0 | 0 | 0 | 0 |
|  | Opacification | Indicator | 24 | -0.0601 | 0.0858 | -0.2219 | 0.0858 |
|  |  |  | 504 | 0.0456 | 0.142 | -0.2254 | 0.179 |
|  | Partial | Indicator | 24 | 0.035 | 0.0281 | 0.0127 | 0.0891 |
|  |  |  | 504 | 0.1562 | 0.0817 | 0.0208 | 0.262 |
| CT Scan | Imaging recorded | Indicator | 24 | 0.0919 | 0.0066 | 0.0066 | 0.1009 |
|  |  |  | 504 | 0.0734 | 0.0133 | 0.0133 | 0.0977 |
|  | Closed Loop Obstruction | Indicator | 24 | 0 | 0 | 0 | 0 |
|  |  |  | 504 | 0 | 0 | 0 | 0 |
|  | Early Or Partial | Indicator | 24 | 0 | 0 | 0 | 0 |
|  |  |  | 504 | 0 | 0 | 0 | 0 |
|  | Ileus | Indicator | 24 | -0.104 | 0.0034 | -0.1099 | 0.0034 |
|  |  |  | 504 | -0.1332 | 0.0139 | -0.1474 | 0.0139 |
|  | Impression High Grade | Indicator | 24 | 0 | 0 | 0 | 0 |
|  |  |  | 504 | 0 | 0 | 0 | 0 |
|  | Internal Hernia | Indicator | 24 | 0 | 0 | 0 | 0 |
|  |  |  | 504 | 0 | 0 | 0 | 0 |
|  | Ischemia | Indicator | 24 | 0.307 | 0.0237 | 0.0237 | 0.3537 |
|  |  |  | 504 | 0.0421 | 0.02 | 0.0181 | 0.0689 |
|  | No Ascites | Indicator | 24 | 0 | 0 | 0 | 0 |
|  |  |  | 504 | 0 | 0 | 0 | 0 |
|  | Pneumatosis Or Free | Indicator | 24 | 0 | 0 | 0 | 0 |
|  |  |  | 504 | 0 | 0 | 0 | 0 |
|  | Transition Point | Indicator | 24 | 0 | 0 | 0 | 0 |
|  |  |  | 504 | 0 | 0 | 0 | 0 |
|  | Unchanged | Indicator | 24 | -0.0102 | 0.0104 | -0.028 | 0.0104 |
|  |  |  | 504 | -0.056 | 0.0201 | -0.0875 | 0.0201 |
